# Generalisability of Results from UK Biobank: Comparison With a Pooling of 18 Cohort Studies

**DOI:** 10.1101/19004705

**Authors:** G. David Batty, Catharine R. Gale, Mika Kivimäki, Ian J. Deary, Steven Bell

## Abstract

**Background:** The UK Biobank cohort study has become a much-utilised and influential scientific resource. With a primary goal of understanding disease aetiology, the low response to the original survey of 5.5% has, however, led to debate as to the generalisability of these findings. We therefore compared risk factor–disease estimations in UK Biobank with those from 18 nationally representative studies with conventional response rates.

**Methods:** We used individual-level baseline data from UK Biobank (N=502,655) and a pooling of data from the Health Surveys for England (HSE) and the Scottish Health Surveys (SHS), comprising 18 studies and 89,895 individuals (mean response rate 68%). Both study populations were aged 40-69 years at study induction and linked to national cause-specific mortality registries.

**Findings:** Despite a typically more favourable risk factor profile and lower mortality rates in UK Biobank participants relative to the HSE-SHS consortium, risk factors–endpoints associations were directionally consistent between studies, albeit with some heterogeneity in magnitude. For instance, for cardiovascular disease mortality, the age- and sex-adjusted hazard ratio (95% confidence interval) for ever having smoked cigarettes (versus never) was 2.04 (1.87, 2.24) in UK Biobank and 1.99 (1.78, 2.23) in HSE-SHS, yielding a ratio of hazard ratios close to unity (1.02, 0.88, 1.19; p-value 0.76). For hypertension (versus none), corresponding results were again in same direction but with a lower effect size in UK Biobank (1.89; 1.69, 2.11) than in HSE-SHS (2.56; 2.20, 2.98), producing a ratio of hazard ratios below unity (0.74; 0.62, 0.89; p-value 0.001). A similar pattern of observations were made for risk factors (smoking, obesity, educational attainment, and physical stature) in relation to different cancer presentations and suicide whereby the ratios of hazard ratios ranged from 0.57 (0.40, 0.81) and 1.07 (0.42, 2.74).

**Interpretation:** Despite a low response rate, aetiological findings from UK Biobank appear to be generalisable to England and Scotland.

## Research in context

### Evidence before this study

The primary objective of UK Biobank is to identify risk factors for chronic diseases and injuries of public health importance. That the baseline response rate of 5.5% was an order of magnitude lower than comparable studies has raised concerns about the generalisability of its findings.

### Added value of this study

For the first time to our knowledge, we directly compared risk factor–health outcome associations in UK Biobank with a pooling of individual-level data from 18 nationally-sampled cohort studies drawn from England and Scotland, all of which have conventional response rates (mean 68%). Risk factor–mortality associations were directionally the same as comparator studies with some heterogeneity in magnitude.

### Implications of all the available evidence

Despite more favourable levels of baseline characteristics and disease-specific mortality rates in UK Biobank, aetiological findings from UK Biobank appear to be generalisable to England and Scotland. This suggests that studies with exceptionally low response rates, conducted at scale and with a range of exposures, have scientific value.

## Introduction

Well-designed prospective cohort studies have considerable utility in identifying genetic and environmental risk factors for an array of somatic and psychiatric disorders. In the numerous contexts where randomised controlled trials are not feasible, cohort studies provide the best approximation of causality. While deeply-phenotyped studies have existed for decades, recent technological advances have led to low-cost, high-throughput methods to quantify genetic variation which, together with simultaneously expanding prospects for linkage to electronic health and care registries, have made possible the formulation of cohort studies with the capacity to explore gene-environment combinations at previously unheralded scale. Several countries have either established such national ‘biobanks’,^1,2^ are in the process of data collection,^3-5^ or are planning such an endeavour.^6^

One such leading resource is the UK Biobank, an open-access, prospective cohort study comprising over 500,000 middle- and older-aged people.^7,8^ Since the completion of baseline data collection in 2010, use has been high, with the study already yielding over 800 peer-reviewed publications.^9^ While UK Biobank is rare in its combination of size, and breadth and depth of content, it has an unusually low response to its baseline survey: of over nine million individuals sent an invitation to participate, only 5.5% did so.^10^ This achieved response rate was in part driven by the cost- and time-saving decision not to re-contact undecided individuals. The project came in under-budget and ahead of schedule.^11^

Whereas such an approach is procedurally efficient, the long-held view is that epidemiological studies need to achieve response rates as high as 80% if their findings are to be credible.^12,13^ Vigorous debates about the impact of non-response on estimations of chronic disease determinants in UK Biobank, its primary objective, have ensued.^14-23^ Concerns seem to rest on the assumption that, relative to a more representative study, the recently reported more favourable prevalence of selected risk factors and mortality rates in UK Biobank relative to surveys with a higher response^24^ will necessarily affect the generalisability of findings for risk factor–disease associations. The principal investigators of UK Biobank have consistently maintained that, provided the exposures of interest are sufficiently varied and the sample suitably large, the generalisability of risk factor–health outcome relationships is assured.^11,25,26^

To address this central issue of whether risk factors for health outcomes are generalisable in UK Biobank, we directly compare effect estimates for risk factors known to be linked to major causes of mortality with those from a pooling of raw data from 18 nationally-sampled cohort studies drawn from England and Scotland, all of which have a response proportion within the conventional range.^27,28^ With UK Biobank data being deployed across a range of scientific disciplines, we chose an array of mortality endpoints and exposures. Given the nature of our research question, our focus was not on uncovering new risk factors for these health endpoints, rather, testing risk factor–endpoint associations that are well-established based on very strong observational and/or experimental evidence. We therefore examined demographic, social, behavioural, and biomedical risk factors for cardiovascular disease,^29,30^ in addition to physical stature in relation to cardiovascular disease and cancer,^31-33^ and educational attainment and suicide risk.^34-37^

## Methods

We used individual-level data from both UK Biobank, a prospective cohort study, and a pooling of eighteen cohort studies with identical core protocols – the Health Survey for England (HSE; 15 studies) and the Scottish Health Surveys (SHS; 3 studies) (hereafter, HSE-SHS).^28,38,39^ The sampling and procedures of these studies have been well described.^7,40^ In brief, in UK Biobank, baseline data collection took place between 2006 and 2010 in 22 research assessment centres across the UK, resulting in a sample of 502,655 people aged 40 to 69 years (response rate 5.5%). In HSE and SHS, a total of 193,842 people aged 16-102 years (response rate 68%; range 58-93^41^) participated in home-based data collection between 1994 and 2008.^40^ Analyses of HSE-SHS were, however, limited to 89,895 people (48,364 women) to match the baseline age range in UK Biobank. In UK Biobank, ethical approval was received from the North-West Multi-centre Research Ethics Committee, and the research was carried out in accordance with the Declaration of Helsinki of the World Medical Association. In HSE-SHS, ethical approval for data collection was granted by the London Research Ethics Council, or local research ethics councils. Participants in both studies gave informed consent.

### Assessment of baseline characteristics

In both UK Biobank and HSE-SHS, physician-diagnosis of chronic disease (diabetes, hypertension, cardiovascular disease), use of multivitamins, lipid-, blood glucose-, and blood pressure-lowering drugs, educational attainment, cohabitation status, and cigarette smoking habit were self-reported based on identical or near-identical enquiries. Although physical activity and alcohol intake were collected using somewhat different questions, we harmonised data across studies and derived comparable binary categories (current non-drinker versus the rest; physically inactive versus the rest).

During medical examinations, waist and hip circumference, height and weight were measured directly using standard protocols. Elevated waist: hip ratio was denoted by ≥0.90 for men and ≥0.85 for women;^42^ obesity was indicated by a body mass index of ≥30kg/m^2^.^43^ Forced expiratory volume in one second, a measure of pulmonary function, was quantified using spirometry with the best of three (UK Biobank) or five (HSE-SHS) technically satisfactory blows used. In UK Biobank, seated systolic and diastolic blood pressure measurements were made twice using the Omron HEM-7015IT digital blood pressure monitor (Omron Healthcare)^20^ or, exceptionally, a manual sphygmomanometer (6652 people). An average of the two readings was used herein. In HSE-SHS, three readings were taken using the Dinamap 8100 automated device,^44^ and a mean of the second and third values was used. We defined hypertension according to existing guidelines as systolic/diastolic ≥140/90 mmHg and/or use of antihypertensive medication.^45^ Non-fasting venous blood was drawn in both studies.^46,47^ Assaying took place at dedicated central laboratories for C-reactive protein, glycated haemoglobin, and total and high-density lipoprotein cholesterol.^40,46^

### Ascertainment of cause-specific mortality

Participants in both studies were linked to mortality records using the procedures of the UK National Health Service Central Registry. Underlying cause of death, coded according to the tenth revision of the International Classification of Disease, was extracted from death certificate data.^48^ We generated the following mortality outcomes: cardiovascular disease, all cancers combined, lung cancer, smoking– attributable cancers, obesity-attributable cancers, and suicide. The ICD codes denoting these causes of death are given in supplemental table 1.

### Statistical analyses

Hazard ratios and accompanying 95% confidence intervals were computed using Cox regression models^49^ and adjusted for age and sex. In these survival analyses we censored individuals according to the date of death or the end of follow-up (14^th^ February 2011 in HSE, 31^st^ December 2009 in SHS; 22^nd^ February 2016 for UK Biobank) – whichever came first. To quantify the difference between the hazard ratios in each of the two studies we computed a ratio of the hazard ratio as we have in other contexts^48^ (UK Biobank was the referent), and calculated the p-value for difference using Fisher’s Z score, with Z calculated as the difference between the logarithms of the two study hazard ratios, divided by the square root of the sum of their variances where Z follows a normal distribution (Z=[β1−β2]/√[SE1^2^+SE2^2^]).^50^ Analyses were conducted using Stata version 15.

### Role of the funding source

The funders of the study had no role in study design, data collection, data analysis, data interpretation, or writing of the report. CRG had full access to UK Biobank data and SB had full access to HSE-SHS data. GDB takes responsibility for the decision to submit for publication.

## Results

In **table 1** (biomedical factors) and **supplemental figure 1** (demographic, social, and behavioural factors) we compare the baseline characteristics of participants in UK Biobank with those in the pooling of the 18 HSE-SHS cohorts. UK Biobank study members were less likely to have had a below-university level education, to be living alone or unmarried, to be sedentary, have existing cardiovascular disease, or to be taking pharmacological treatments for raised blood glucose, although the reverse was seen for lipid- and blood pressure-lowering medications. In analyses restricted to study members not reporting the use of such therapies, there was essentially no difference between studies members for total and high-density lipoprotein cholesterol, and glycosylated haemoglobin. Whereas values for C-reactive protein were marginally lower in UK Biobank, both systolic and diastolic blood pressure were somewhat higher. Taken together, there was a generally more favourable risk factor profile in UK Biobank study members.

**Table 1.** Summary of baseline biomedical characteristics in UK Biobank and the HSE-SHS cohort studies.

|  | **UKBB** | **HSE-SHS** |
| --- | --- | --- |
| Number of studies | 1 | 18 |
| Number of adults (women) | 502,655<br>(273,472) | 89,895<br>(48,364) |
| Age, mean (SD), years | 56.5 (8.10) | 53.5 (8.6) |
| FEV1, mean (SD), litres | 2.81 (0.80) | 2.89 (0.89) |
| Total cholesterol, mean (SD), mmol/L | 5.89 (1.07) | 5.95 (1.14) |
| High-density lipoprotein cholesterol, median (IQR), mmol/L | 1.43<br>(1.20-1.71) | 1.40<br>(1.20-1.70) |
| Glycosylated haemoglobin, median (IQR), mmol/mol | 35.0<br>(32.6-37.4) | 36.6<br>(33.3-40.9) |
| C-reactive protein, median (IQR), mg/L | 1.26<br>(0.63, 2.49) | 1.50<br>(0.70, 3.10) |
| Systolic blood pressure, mean (SD), mmHg | 137.7 (19.3) | 133.3 (18.4) |
| Diastolic blood pressure, mean (SD), mmHg | 81.6 (10.6) | 76.6 (11.5) |

In both studies, the sample size is the maximum number with baseline data; numbers are lower for selected variables. Analyses for total cholesterol and high-density lipoprotein excludes participants taking lipid-lowering medication; analyses for glycosylated haemoglobin excludes self-reported diabetics and people taking blood glucose-lowering medication; analyses for C-reactive protein excludes people with values >10 mg/L; and analyses for blood pressure excludes people taking blood pressure-lowering medication. FEV1, forced expiratory volume in one second; IQR, interquartile range; and SD, standard deviation.

In UK Biobank, there were 14,288 deaths during an average of 7.0 years of follow up in 499,701 people consenting to be linked to mortality registers. In the pooling of data from HSE-SHS, 10 years of mortality surveillance gave rise to 7,861 deaths in 89,895 individuals. Of the five mortality categories examined, there were markedly lower rates of cardiovascular disease, all cancers combined, and tobacco- and obesity-attributable cancers in UK Biobank, whereas the rate of suicide was higher (supplemental Table 2).

In **figure 1**, for each dataset, we depict the direction and strength of the relationship of baseline demographic and behavioural characteristics known to be associated with cardiovascular disease mortality. All well-established risk factors for cardiovascular disease were recapitulated in the present analyses of both studies. The expected direction of association was the same in both studies for the seven characteristics, whereby being male, of higher age, being physically inactive, a non-drinker of alcohol, not being married/cohabiting, a current or former smoker, and not having a higher education degree were related to elevated rates of cardiovascular disease mortality. There was, however, a difference in the magnitude of these effects in four of the risk factors examined, and in most of these the hazards ratios were higher in UK Biobank. When we explored the links between biomedical factors and cardiovascular disease mortality (**figure 2**), again, all ten of the biomarkers studied revealed known associations with cardiovascular disease deaths in both studies. There was also some heterogeneity in the strength of these relationships for higher levels of glycosylated haemoglobin, existing cardiovascular disease (stronger effects in UK Biobank than HSE-SHS for both risk factors) and hypertension (the reverse).

**Figure 1.**
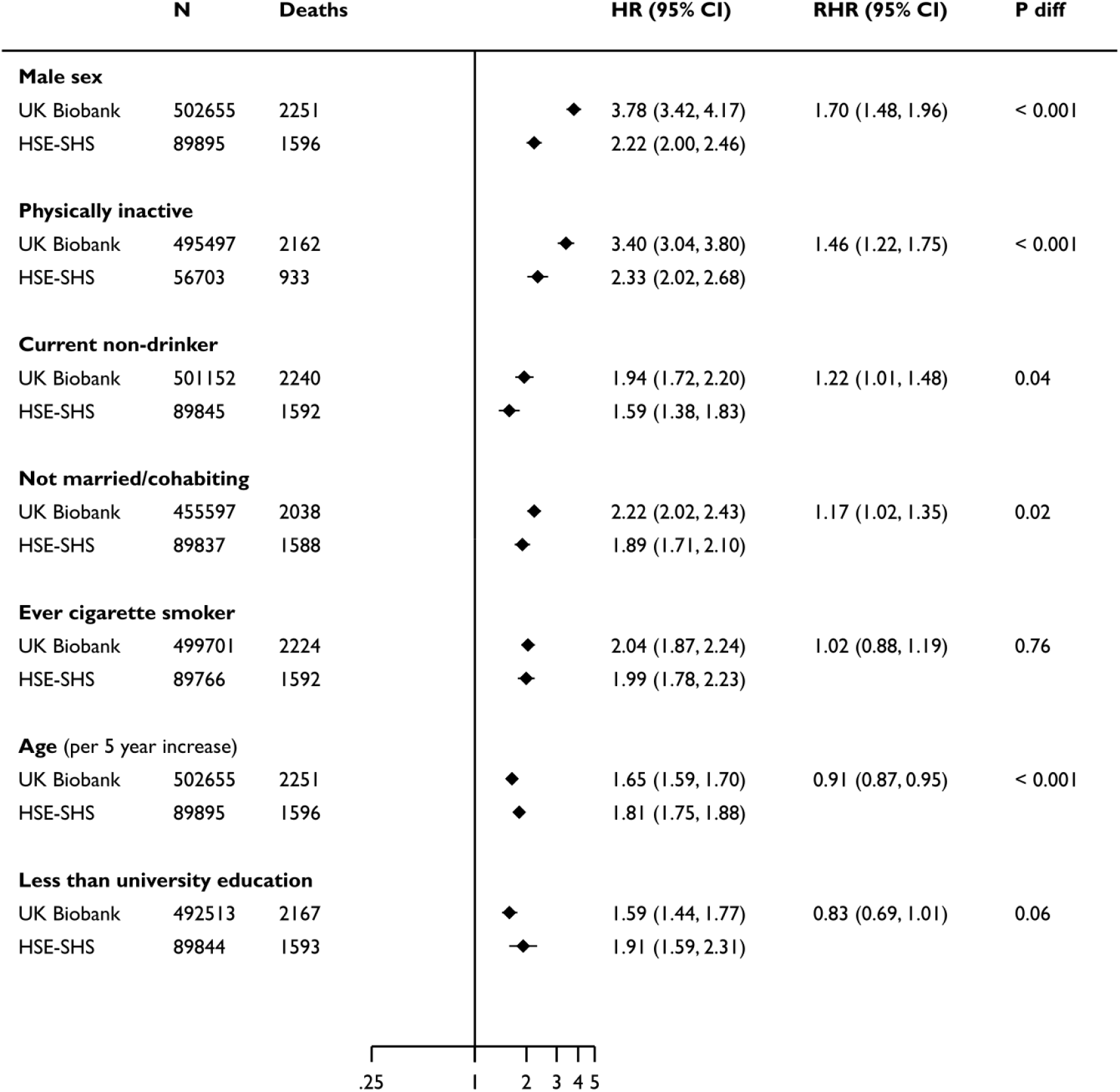
Association of baseline demographic and behavioural characteristics with cardiovascular disease mortality in UK Biobank and the HSE-SHS cohort studies. Hazard ratios are age- and sex-adjusted with the exception of age and sex which are mutually adjusted. The shaded diamonds indicate the hazard ratio (HR) and error bars denote the 95% confidence interval (CI) for the relation of each characteristics with the risk of death outcome. The ratio of hazard ratio (RHR) summarises the difference (HSE-SHS is the referent group) between that effect estimates for the outcome, and the p-value is to determine if the observed differences between groups are significant. In both studies, the sample size is the maximum number with baseline data; numbers are lower for selected variables.

**Figure 2.**
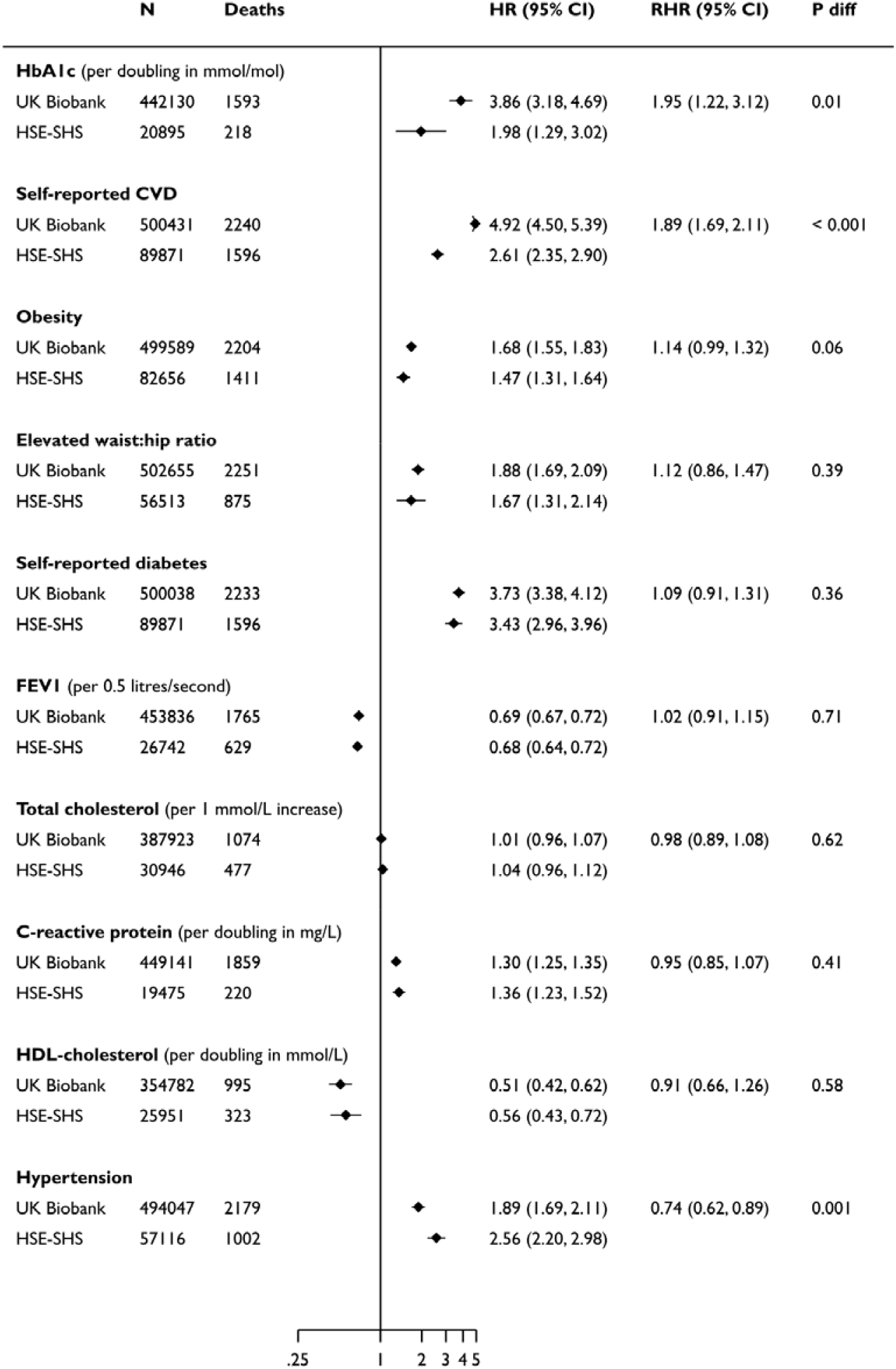
Association of baseline biomedical characteristics with cardiovascular disease mortality in UK Biobank and the HSE-SHS cohort studies. Hazard ratios are age- and sex-adjusted. The shaded diamonds indicate the hazard ratio (HR) and error bars denote the 95% confidence interval (CI) for the relation of each characteristics with the risk of death outcome. The ratio of hazard ratio (RHR) summarises the difference (HSE-SHS is the referent group) between that effect estimates for the outcome, and the p-value is to determine if the observed differences between groups are significant. The distributions of glycosylated haemoglobin (HbA1c), C-reactive protein, and HDL-cholesterol were skewed, hence they were log_2_-transformed and effect estimates reflect a doubling in the biomarker. Elevated waist: hip ratio was denoted by ≥0.90 for men and ≥0.85 for women; obesity was indicated by a body mass index of ≥30 kg/m^2^. CVD, cardiovascular disease; FEV1, forced expiratory volume in one second; HDL, high-density lipoprotein. In both studies, the sample size is the maximum number with baseline data; numbers are lower for selected variables

Nest, we examined the association of selected baseline factors with some non-cardiovascular mortality outcomes, including different presentations of cancer and also suicide deaths (**figure 3**). These known risk factors were replicated across both studies. The magnitude of the association of cigarette smoking with lung cancer and malignancies aetiologically linked with tobacco were weaker for UK Biobank, whereas obesity and obesity-attributed cancers yielded similar associations in each study. Hazard ratios were also essentially the same for lower educational attainment and suicide although statistical power was modest in these analyses as evidenced by the wide confidence intervals, particularly for HSE-SHS. Physical stature revealed the predicted opposing and shallow gradients for cardiovascular disease (negative) and cancer (positive); again, effect sizes were very similar in both studies.

**Figure 3.**
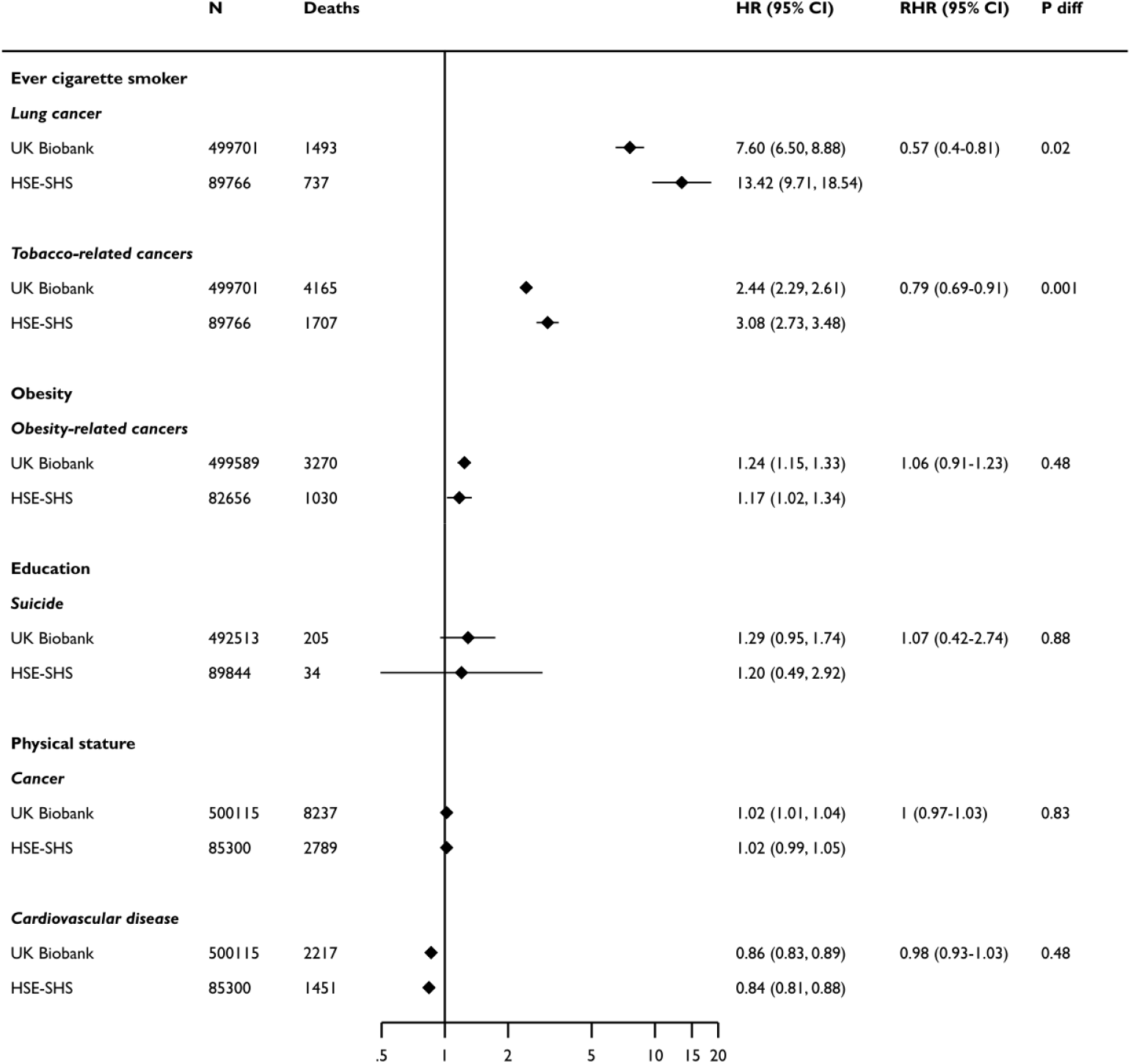
Association of selected baseline characteristics with cause-specific mortality in UK Biobank and the HSE-SHS cohort studies. Hazard ratios are age- and sex-adjusted. The shaded diamonds indicate the hazard ratio (HR) and error bars denote the 95% confidence interval (CI) for the relation of each characteristics with the risk of death outcome. The ratio of hazard ratio (RHR) summarises the difference (HSE-SHS is the referent group) between that effect estimates for the outcome, and the p-value is to determine if the observed differences between groups are significant. Analyses for education were based on less than university education versus the rest; a body mass index of ≥30 kg/m^2^ for obesity; and a per 5 cm increase in height. In both studies, the sample size is the maximum number with baseline data; numbers are lower for selected variables.

In sensitivity analyses, we explored the impact of using the same calendar period of study induction in both UK Biobank and HSE-SHS – whereas baseline in UK Biobank was in fact 2006-2010, the latest survey for which we have linked data in the HSE-SHS collaboration was 2008. Results of these subgroup analyses had little impact on our overall conclusions (**supplemental figure 2**). The only obvious anomaly was for obesity which might be explained by the small number of cardiovascular disease deaths. We also examined if results from survival analyses featuring cardiovascular disease as our outcome of interest differed across studies in gender-specific analyses for demographic and behavioural characteristics (**supplemental figure 3**), and biomarkers (**supplemental figure 4**). There was no suggestion of this being the case. Lastly, given that there was, as described, a higher use of medication for blood pressure- and lipid-lowering in members of UK Biobank relative to our comparator cohorts, we tested if this was also evident for other health-seeking behaviours such as vitamin and mineral supplementation. The prevalence of such use was in fact lower in UK Biobank (21.8%) than HSE-SHS (33.1%).

## Discussion

Our main finding was that in a comparison of findings between UK Biobank and a pooling of 18 studies from the HSE-SHS consortium, well-established risk factor–mortality associations were recapitulated. While these effects estimates were directionally consistent between studies there was some difference in magnitude. This general similarity of results was apparent despite the response rate in UK Biobank being an order of magnitude lower than comparator cohorts, and UK Biobank members having a generally more favourable prevalence of sociodemographic, behavioural, and health characteristics at baseline, and lower rates of cause-specific mortality during follow-up.

### Findings from other studies

The only other analyses of risk factor–disease relationships in UK Biobank against comparison studies of which we are aware is that for venous thromboembolism in the Emerging Risk factors Collaboration, a pooling of data from 76 cohort studies.^51^ The goal of that paper was risk factor discovery as opposed to testing well-established associations between risk factors and chronic disease. Inter-study comparison was further hampered by differing endpoint ascertainment and blood-based biomarkers from UK Biobank not being available at the time of analyses.

As described, UK Biobank investigators, while acknowledging their study has little value in describing the current prevalence of a risk factor or a disorder – never a stated objective – have attempted to minimise unease around the investigation of chronic disease aetiology – its primary purpose – by arguing that generalisable associations with risk factors can be obtained in non-representative samples provided there are sufficiently large numbers of individuals with a range of exposures.^11,25,26^ They cite, as circumstantial evidence, examples of cohort studies drawing on highly selected populations that have markedly high response rates than UK Biobank – Framingham residents,^52^ British physicians,^53^ US nurses^54^ – all of which produced results that have subsequently been shown to be transportable to general population-based studies and have contributed much to the prevention of cardiovascular disease and selected cancers. Similarly, our present findings mirror those from analyses where we have compared risk factors for coronary heart disease in another very select group, a cohort of British civil servants (the Whitehall II prospective cohort study), with those from a cohort based on the general population (the British Regional Heart Study).^55^ In those analyses we found near-identical risk factor–disease association across studies.

### Strengths and limitations

The strengths of the present study include the comparison of UK Biobank with well-characterised and geographical representative studies that have high response rates. Our report inevitably has some limitations, however. First, whereas UK Biobank includes people from the contiguous countries that comprise the UK, there were no data from Wales in the comparator studies. Second, whereas core elements of data collection in the HSE-SHS consortium were constant across studies, scientific themes for data collection differed from year to year.^40^ As such, selected biomedical data were not collected in all cohorts and the analytical sample size was therefore lower for selected analyses. Third, for two variables – physical activity and alcohol intake – baseline data were not directly comparable between studies, though we were able to harmonise data into binary groups. Lastly, although blood samples have been frozen in HSE-SHS, so raising the potential for later genome sequencing, comparison with genetic risk prediction of chronic disease in UK Biobank are currently not possible.

In conclusion, the finding that UK Biobank reveals similar risk factor–mortality associations to those evident in comparator studies with conventional response rates would appear to challenge the prevailing view of a lack of generalisability of aetiological findings in this cohort. This suggests that the cost- and time-savings resulting from its low response rate did not impact upon scientific utility.

## Data Availability

Data from The UK Biobank (http://www.ukbiobank.ac.uk/), and The Health Surveys for England and The Scottish Health Surveys (https://data-archive.ac.uk/) are available to researchers upon application.

## Acknowledgements

GDB is supported by the UK Medical Research Council (MR/P023444/1) and the US National Institute on Aging (1R56AG052519-01; 1R01AG052519-01A1); MK by the UK Medical Research Council (MR/R024227), US National Institute on Aging (NIH), US (R01AG056477), NordForsk, and Academy of Finland (311492); CRG by the UK Medical Research Council (MRC_MC_UU_12011/2 and MRC_MC_UP_A620_1015); and SB by the NIHR Blood and Transplant Research Unit in Donor Health and Genomics (NIHR BTRU-2014-10024), UK Medical Research Council (MR/L003120/1), British Heart Foundation (RG/13/13/30194), and NIHR Cambridge Biomedical Research Centre at the Cambridge University Hospitals NHS Foundation Trust. There was no direct financial or material support for the work reported in the manuscript.

## Access to data

Data from The UK Biobank (http://www.ukbiobank.ac.uk/), and The Health Surveys for England and The Scottish Health Surveys (https://data-archive.ac.uk/) are available to researchers upon application. Part of this research has been conducted using the UK Biobank Resource under Application 10279.

## Conflict of interest

All authors are registered and enthusiastic users of UK Biobank. Ian Deary had a role in the design of the cognitive function battery.

## Contributions

GDB generated the idea for the present paper, formulated an analytical plan, and wrote the manuscript; CRG (UK Biobank) and SB (HSE-SHS) formulated an analytical plan and carried out the data analyses; and SB prepared the figures. All authors commented on an earlier version of the manuscript.

---

**Supplemental Table 1.** Mortality outcomes in the present analyses.

| **Mortality outcome** | **ICD 10 codes** |
| --- | --- |
| All cancers combined | C00-C97 |
| Obesity-attributable cancers<sup>56</sup> | Breast (postmenopausal) C50; Colon C18; Rectum C19-20; Pancreas C25; Endometrium C54; Kidney C64; Ovary C56 & C57.0-4; Multiple myeloma C90; Liver C22; Thyroid C73; Gallbladder C23-24; Oesophageal adenocarcinoma C15 & morphology 8140-8141, 8143-8145, 8190-8231, 8260-8263, 8310,8401, 8480-8490, 8550-8551, 8570-8574, 8576; and Gastric cardia C16.0 |
| Smoking-attributable cancers<sup>57,58</sup> | Lip, oral cavity, pharynx C00-C14, oesophagus C15, stomach C16, colon and rectum C18-C20, liver C22, pancreas C25, larynx C32, trachea, lung, bronchus C33-C34, cervix uteri C53, kidney and renal pelvis C64-C65, urinary bladder C67, and acute myeloid leukemia C92.0 |
| Cardiovascular disease | I20-5, I50, I60-70, I73, I74 |
| Suicide<sup>34</sup> | Suicide and self-inflicted poisoning by solid or liquid substances (E950-E959), injury undetermined whether accidentally or purposely inflicted (E980-E989), terrorism (U03.1 and U03.9), intentional self-harm (X60-X84), event of undetermined intent (Y10-Y34), sequelae of intentional self-harm, assault and events of undetermined intent (Y87), and sequelae of unspecified external cause (Y89.9)) |
| Lung cancer | C34 |

**Supplemental Table 2.** Mortality rates in participants in UK Biobank and the HSE-SHS cohort studies.

|  | **UKBB** | **HSE-SHS** |
| --- | --- | --- |
| Number of adults (women) | 502,655<br>(273,472) | 89,895<br>(48,364) |
| Duration of mortality surveillance (years, mean [SD]) | 6.99 (1.03) | 9.62 (4.39) |
| Total number of deaths | 14,288 | 7,861 |
| Mortality rate for cardiovascular disease | 64 | 185 |
| Mortality rate for all cancers combined | 236 | 342 |
| Mortality rate for tobacco-attributable cancers | 120 | 198 |
| Mortality rate for obesity-attributable cancers | 94 | 132 |
| Mortality rate for suicide | 6 | 4 |

Mortality rates are per 100,000 person-years. Analyses are based on 502,655 people in UK Biobank and 89,895 in HSE-SHS. SD, standard deviation.

**Supplemental Figure 1.**
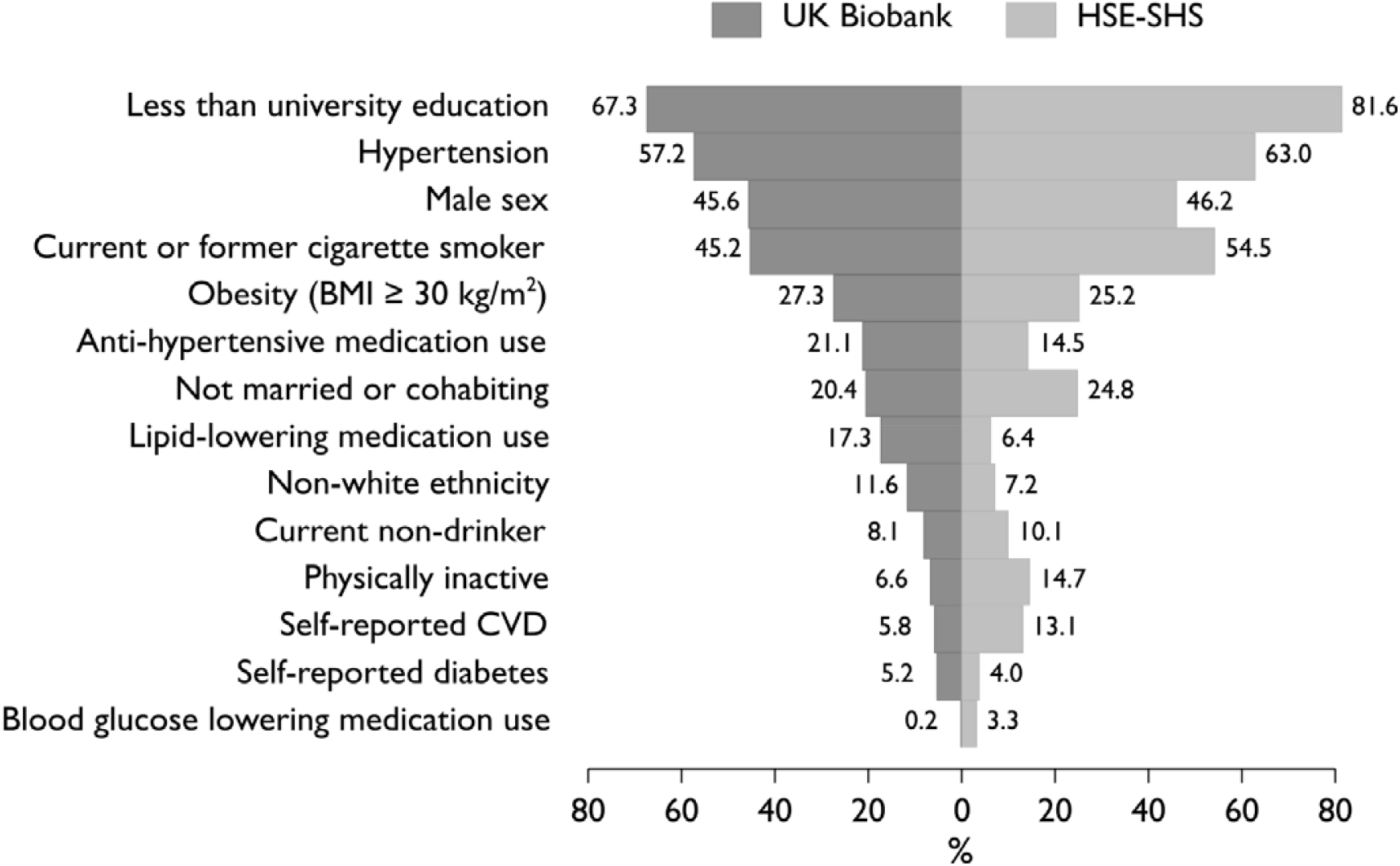
Prevalence of baseline characteristics in UK Biobank and the HSE-SHS cohort studies*. *Selected comparisons have been published by Fry and others^24^ using published results from four HSE studies (2006, 2008, 2009, and 2010). Our analyses are based on analyses of raw data from 18 cohort studies from England and Scotland which include two of those HSE studies (2006, 2008). Analyses comprise 502,655 people in UK Biobank and 89,895 in HSE-SHS. BMI, body mass index; CVD, cardiovascular disease.

**Supplemental Figure 2.**
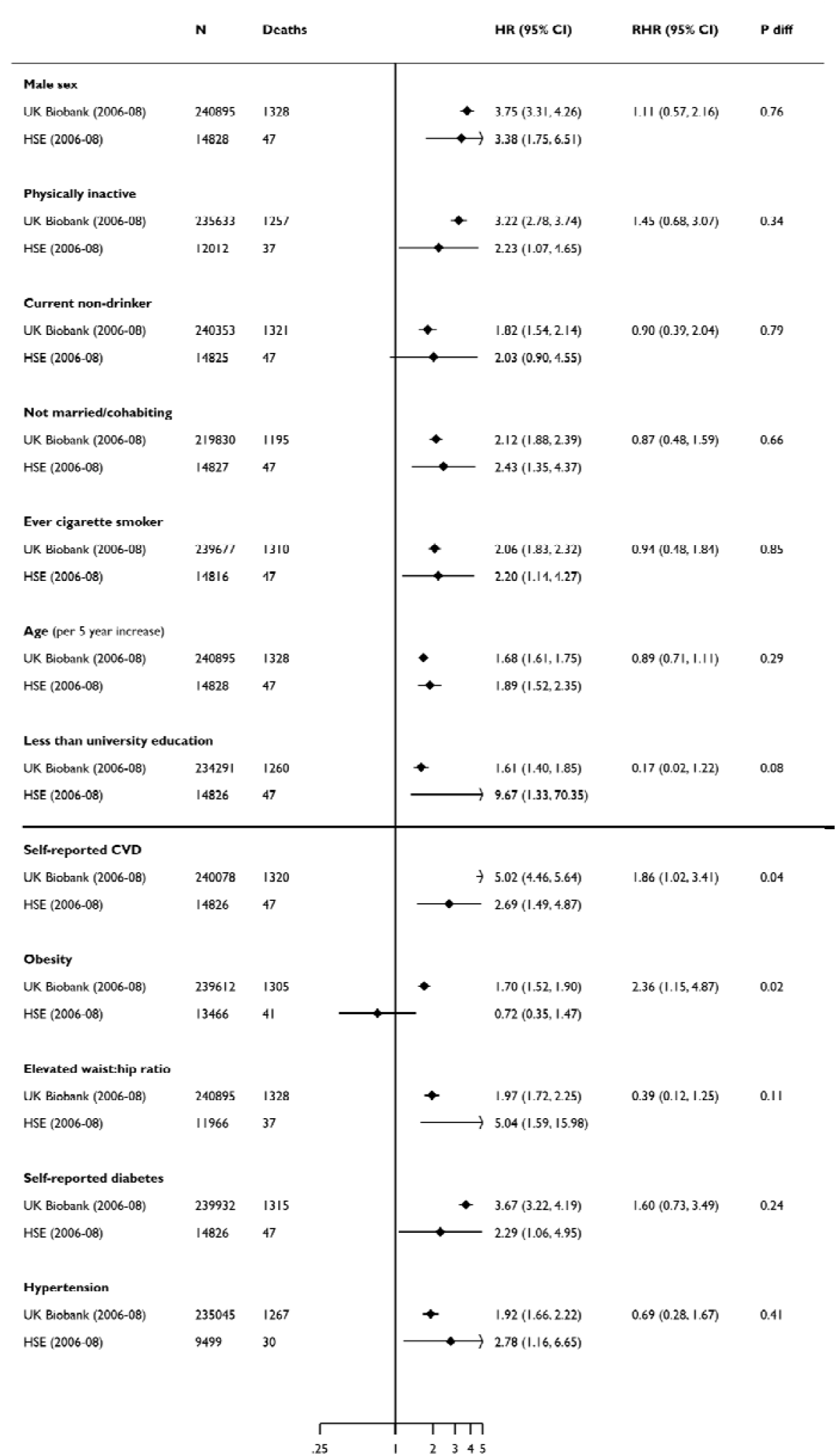
Association of selected baseline characteristics with cardiovascular disease mortality in UK Biobank and the HSE-SHS cohort studies – analyses based on the same survey years (2006-08) Hazard ratios are age- and sex-adjusted with the exception of age and sex which are mutually adjusted. The shaded diamonds indicate the hazard ratio (HR) and error bars denote the 95% confidence interval (CI) for the relation of each characteristics with the risk of death outcome. The ratio of hazard ratio (RHR) summarises the difference (HSE-SHS is the referent group) between that effect estimates for the outcome, and the p-value is to determine if the observed differences between groups are significant. Elevated waist: hip ratio was denoted by ≥0.90 for men and ≥0.85 for women; obesity was indicated by a body mass index of ≥30 kg/m^2^. CVD, cardiovascular disease. In both studies, the sample size is the maximum number with baseline data; numbers are lower for selected variables.

**Supplemental Figure 3.**
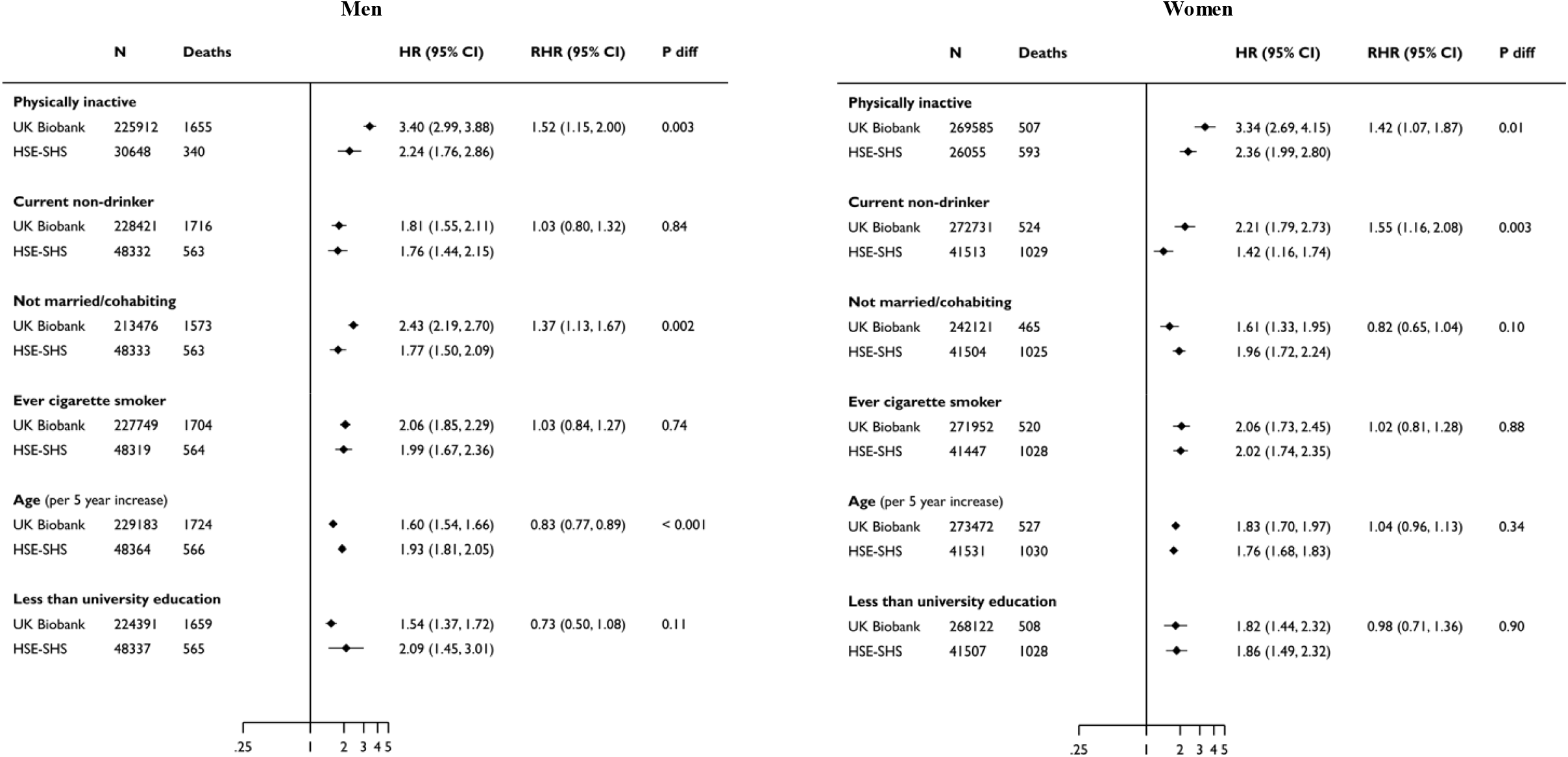
Association of baseline demographic and behavioural characteristics with cardiovascular disease mortality in UK Biobank and the HSE-SHS cohort studies – stratification by gender. Hazard ratios are age- and sex-adjusted. The shaded diamonds indicate the hazard ratio (HR) and error bars denote the 95% confidence interval (CI) for the relation of each characteristics with the risk of death outcome. The ratio of hazard ratio (RHR) summarises the difference (HSE-SHS is the referent group) between that effect estimates for the outcome, and the p-value is to determine if the observed differences between groups are significant. In both studies, the sample size is the maximum number with baseline data; numbers are lower for selected variables.

**Supplemental Figure 4.**
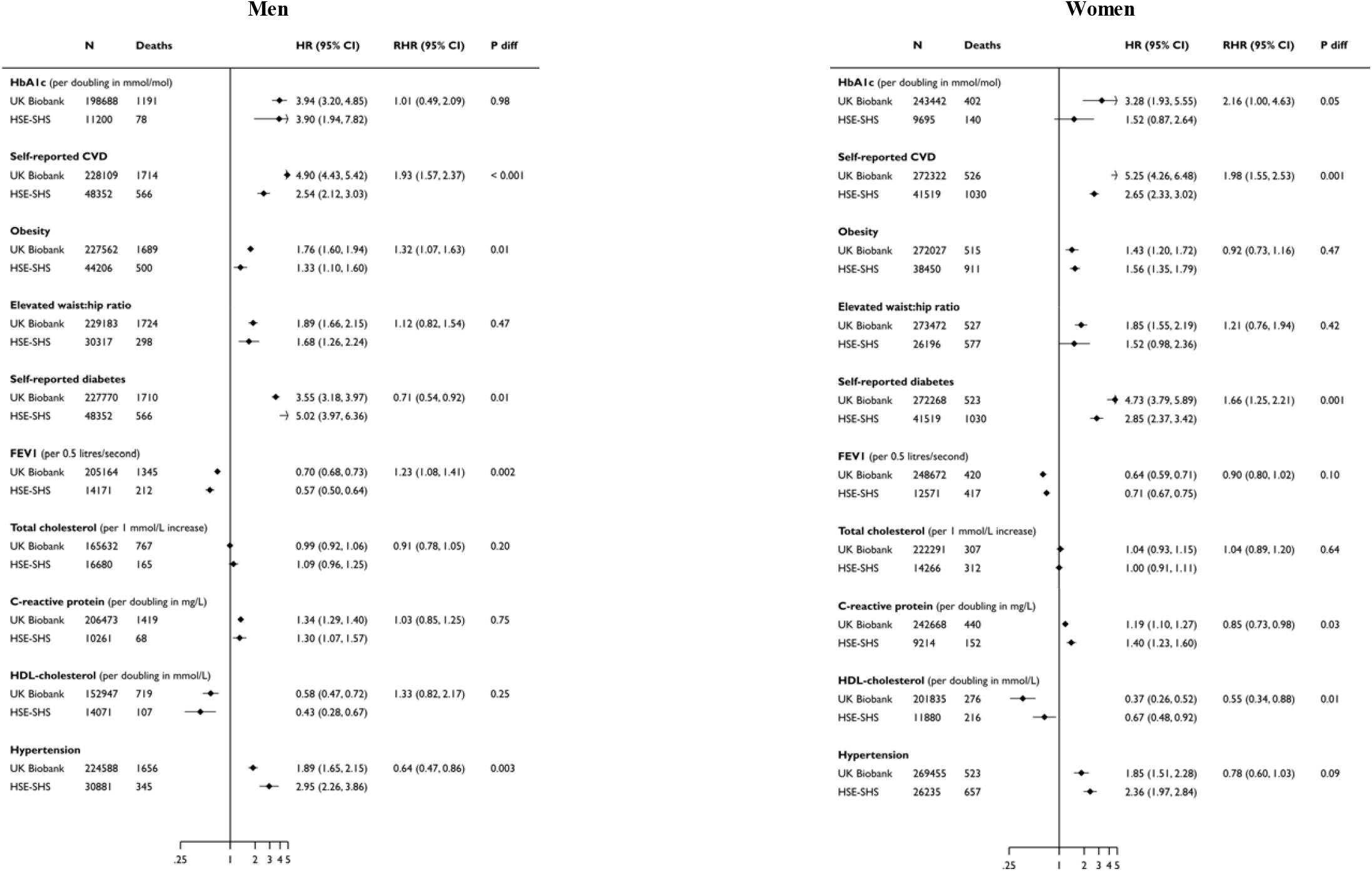
Association of baseline biomedical characteristics with cardiovascular disease mortality in UK Biobank and the HSE-SHS cohort studies – stratification by gender. Hazard ratios are age- and sex-adjusted. The shaded diamonds indicate the hazard ratio (HR) and error bars denote the 95% confidence interval (CI) for the relation of each characteristics with the risk of death outcome. The ratio of hazard ratio (RHR) summarises the difference (HSE-SHS is the referent group) between that effect estimates for the outcome, and the p-value is to determine if the observed differences between groups are significant. HbA1c, glycosylated haemoglobin; FEV1, forced expiratory volume in one second; and HDL, high-density lipoprotein. In both studies, the sample size is the maximum number with baseline data; numbers are lower for selected variables.

## Notes

### Competing Interest Statement

The authors have declared no competing interest.

### Author Declarations

All relevant ethical guidelines have been followed and any necessary IRB and/or ethics committee approvals have been obtained.

Any clinical trials involved have been registered with an ICMJE-approved registry such as ClinicalTrials.gov and the trial ID is included in the manuscript.

